# Optimized hypertension care for people with high blood pressure by improved integrated care and self-management tools: a mixed-methods study

**DOI:** 10.64898/2026.05.14.26352728

**Authors:** Saskia E. van Grondelle, Annefrans F.T.M. van Ede, Jonne G. ter Braake, Sytske van Bruggen, Guy E.H.M. Rutten, Michiel L. Bots, Hedwig Vos, Mattijs E. Numans, Rimke C. Vos

## Abstract

**Background:** High blood pressure (BP) is an often treatable cause of cardiovascular disease. We developed an intervention, featuring a cardiovascular expert team and a toolbox, to support healthcare professionals (HCPs) in hypertension management and enhance patient self-management.

**Aim:** This study evaluates the adoption and feasibility of this intervention.

**Design and Setting:** A mixed-methods study in general practices in the Netherlands.

**Methods:** HCPs could consult a cardiovascular expert team and use a self-management toolbox for their patients as preferred. We interviewed HCPs guided by the Consolidated Framework of Implementation Research (CFIR), and HCPs completed the Determinants of Implementation Behaviour Questionnaire (DIBQ). Using CFIR-ERIC matching tool, we matched implementation strategies to identified barriers. Adults with elevated BP, who were prescribed at least two blood pressure lowering medications were eligible to participate. Patient and disease characteristics were extracted from the electronical medical record.

**Results:** Of 591 eligible patients at thirteen general practices, 176 participated. The cardiovascular expert team was well-received, with 33 unique consultations, although nurse practitioners (NPs) might need the expertise of the expert team more frequently than general practitioners (GP) (adoption). The toolbox was perceived as challenging to use (feasibility). We subsequently identified three key strategies to improve implementation. Mean systolic and diastolic BP were 158/87 mmHg at baseline and 148/85 mmHg after 12 months, although this change cannot be conclusively linked to the intervention.

**Conclusions:** Structured implementation strategies may be helpful in hypertension management. The cardiovascular expert team was considered valuable, but might be better targeted to NPs rather than GPs.

## Introduction

High blood pressure (BP) or hypertension (repeated systolic blood pressure > 140 mmHg) is associated with increased cardiovascular morbidity and mortality(1), but risk can be reduced through lifestyle interventions and blood pressure-lowering medications(2). Nonetheless, control of hypertension remains a worldwide challenge, and in high-income countries only around 40% of individuals with hypertension achieves their target BP(1, 3). This results in a significant burden on both individuals and healthcare systems.

Hypertension which does not respond to lifestyle change or BP-lowering medications can be caused by resistant hypertension, which is defined as a systolic BP > 140 mmHg or a diastolic BP > 90 mmHg when prescribed three or more antihypertensive medications, including a diuretic. This form of hypertension has a poor prognosis and is a reason for referral to secondary care. However, most cases of uncontrolled hypertension are due to modifiable behavioural factors, such as medication non-adherence(4), adverse diet and lifestyle factors such as high salt intake, overweight and smoking(5, 6). Social determinants of health (SDOH), like (financial) stress and environmental factors also affect hypertension control. For example, limited access to healthcare, financial instability, the lack of social support and the stress associated with these conditions can all obstruct effective management of hypertension(7). Behavioural determinants and SDOH are primarily dealt with in primary care, since healthcare providers (HCP) in primary care are relatively more familiar with a patient’s individual context through continuity of care.

For HCPs it is not always easy to distinguish patients truly resistant to pharmacological therapy from those whose BP is uncontrolled due to behavioural factors or SDOH. Avoidable referrals to secondary care (for medical as well as non-medical causes of hypertension) may therefore occur(8). To identify areas of potential improvement, we studied the barriers and facilitators experienced in hypertension treatment. HCPs expressed a need for better collaboration with secondary care, tools to improve interpersonal skills and more time for communication with patients(9). Based on these results we developed a multi-component intervention to improve hypertension care, focused on supporting the HCP in clinical decision making and stimulating self-management in their patients. By supporting HCPs, we hope to promote appropriate care, reduce avoidable referrals to secondary care and aid HCPs in hypertension management. In this study, we investigated the adoption, feasibility and potential beneficial health outcomes of this multi-component intervention.

## Methods

### Setting and study design

Between November 2022 and November 2023, we conducted a convergent mixed-methods implementation study of the adoption and feasibility of a multi-component intervention focused on people with hypertension in general practices. One of the primary components of the intervention was a cardiovascular expert team, offering easily accessible and tailored advice on whether to focus on behavioural factors or SDOH, adjust pharmacological treatment, or initiate referral to secondary care due to suspected secondary hypertension. In addition, the intervention consisted of a toolbox to stimulate self-management and improve lifestyle, consisting of BP monitors, dialogue tools and pedometers (Supplementary Box 1). We also assessed preliminary improvements of health outcomes. Adoption was defined as ‘the intention, initial decision to try the multi-component intervention’ and feasibility as ‘the extent to which the multi-component intervention, can be successfully used in primary care’(10). Outcomes were assessed via interviews with HCPs and an online survey to understand perception and usage of the intervention. Furthermore, we used the rate of referrals to the expert team as an indicator of the intervention’s impact and implementation. Because implementation research is an iterative process, we collected data during the study period and evaluated adoption, feasibility and preliminary effects on health outcomes at 12 months.

### Inclusion process and criteria

We deployed a stepped inclusion process to recruit people with hypertension in the area of The Hague. First, we identified eligible general practices and included those with HCPs willing to participate. All general practices in the highly urbanized region of The Hague (approximately half a million inhabitants) were eligible for participation, but during inclusion we opted for variation in neighbourhood socio-economic position (SEP) between practices. Adults at the participating practices were eligible if they had an elevated BP and were prescribed at least two BP-lowering medications. Participants provided written consent for use of their routine care data for evaluation.

### Ethical considerations

The quality of the protocol was reviewed and approved by the scientific committee of the department of Public Health and Primary Care (WSC-2020-30/SP). The Medical Ethics Review Board Leiden, The Hague, Delft and provided a declaration of no objection (Protocol number: P20-092) and deemed that our study was not subject to the Dutch Medical Research with Human Subjects Law.

### Data collection

We assessed adoption and feasibility using triangulation of qualitative and quantitative data. We performed semi-structured interviews with participating HCPs and sent out a survey to identify both contextual barriers and facilitators influencing implementation, as well as those of behaviour change during implementation. In addition, the number of referrals to the cardiovascular expert team was assessed using the teleconsultation system. For each referral, HCPs also recorded in the teleconsultation system what action they would have taken without the support of an expert team, such as referral to secondary care, contacting a medical specialist by phone or continuing treatment in primary care. Lastly, biomedical determinants (BP, glucose, HbA1c, LDL, BMI) and age, sex, number and type of medication were extracted from the electronic medical records (EMR).

### Interviews – Consolidated Framework of Implementation Research (CFIR)

To identify barriers and facilitators influencing the adoption and feasibility of the multi-component intervention, semi-structured interviews were conducted and recorded by SG. Each participating practice was contacted to arrange interviews, while the practices themselves determined the timing and number of HCPs involved per interview. The interview guide was developed using the CFIR framework, which is one of the most widely used implementation frameworks to guide assessment of contextual determinants of implementation(11). The CFIR consists of five domains, ‘innovation’, ‘outer setting’, ‘inner setting’, ‘individuals’ and ‘implementation process’, each of which consists in turn of different assessable constructs. For the purpose of this study the interview guide concentrated on intervention implementation and the reasons behind the use or non-use of its components as assessed in the CFIR (Supplementary Box 2).

### Survey - Determinants of Implementation Behaviour Questionnaire (DIBQ)

A modified version of the Determinants of Implementation Behaviour Questionnaire (DIBQ) was used to determine barriers and facilitators of behavioural change of participating HCPs (Supplementary Box 3). The DIBQ is a validated questionnaire based on the theoretical domains framework (TDF)(13). TDF domains represent theory-based factors relevant for implementation behaviours and behavioural change. DIBQ questions measure TDF domains. The survey was sent by e-mail to all participating HCPs, who were given one month to respond, with a reminder sent after two weeks.

### Patient-related characteristics

Patient-related characteristics were extracted from the EMR and pseudonymized by a trusted external healthcare partner, and included systolic blood pressure (SBP) (mmHg), diastolic blood pressure (DBP) (mmHg), (fasting) glucose (mmol/L), HbA1c (mmol/mol), eGFR (ml/min/1.7), LDL (mmol/L), BMI (kg/m^2^), multi-morbidity, use of cardiovascular medication, age and sex. As SDOH are a known risk factor for hypertension, we assessed if participants were living in high or low SEP areas. For this purpose a ‘deprivation score’ was used(14), which indicates the average SEP of the population in an area relative to the entire city. These deprivation scores are based on five indicators: 1) the percentage of individuals with a migration background from non-Western countries; 2) the percentage of individuals receiving social support; 3) the average household income; 4) the average property value, and 5) the percentage of residents who have moved in the past three years in or out of the neighbourhood. A negative score (-) indicates less or no disadvantage, while a positive score indicates more disadvantage. For The Hague, the average score was set at 0. Therefore, a negative or positive score signifies a deviation from the average score of The Hague. We identified negative scores as high SEP and positive scores as low SEP.

### Data analysis

#### Interviews

Interviews were transcribed verbatim and analysed using the Framework Method, which allows systemic data analysis(15). First, we imported all data in Atlas.ti (24) and relevant quotes were highlighted by researchers. The quotes were then categorized into CFIR constructs using a deductive approach. The first two interviews were coded independently by SG and AE, after which the codes were discussed until consensus was reached. SG coded the remaining interviews and discussed them with AE. An additional meeting with SG, AE and RV was organized to discuss interpretation of data and ensure trustworthiness of results. The final matrix consisting of summaries per construct of the CFIR was assessed by SG and RV. Then, the most important barriers per construct were identified and we matched implementation strategies to the selected barriers and facilitators using CFIR’s Expert Recommendations for Implementing Change (ERIC) matching tool(16). This tool supports formulating context-fitting strategies to overcome barriers and accentuate facilitators in implementation processes. For each CFIR barrier, the ERIC matching tool provides corresponding strategies to enhance implementation. The final strategies that could potentially increase intervention adoption were discussed and chosen in an additional meeting with SG, RV and AE.

#### Survey

Data analyses were performed in R studio version 22.07.01. Each DIBQ answer ranges from 1 to 5. For most questions, a low score indicates a potential barrier to implementation behaviour within a domain, while a high score suggests that the domain facilitates implementation. For questions 21, 22, and 33, the interpretation is reversed and these questions were therefore reverse coded before analysis. Means and standard deviations were calculated per question and per domain.

#### Patient-related characteristics

We used descriptive statistics in R studio version 22.07.01 for analysis. We compared mean BP, number of prescribed BP-lowering medications, eGFR, LDL and BMI as recorded in the year before the study to the mean of all available measurements taken during follow-up, stratifying by SEP area. Due to the use of routine care data, the extent of missing data varied. We therefore conducted a complete case analysis to evaluate whether missing data impacted the results.

## Results

We included 13 out of 18 eligible practices in The Hague distributed across high SEP (n=6) and low SEP (n=7) areas, which provide care to up to 48,719 patients. Mean age of HCPs was 48 years (8 sd) and 73% were women. The total number of general practitioners (GP) working in the practices was 32, with an average of 2.5 GPs per practice (1.8 sd). The total number of NPs working in the practices was 16, with an average of 1.2 NPs (0.7 sd) per practice. Of these HCP, we interviewed 7 GPs and 5 NPs (the practice-based contact persons) from 12 practices. Interviews lasted approximately 30 minutes and were digitally recorded.

### Barriers

Three main barriers were identified. The first was a lack of perceived urgency to change existing workflows, which was linked to different constructs within the ‘inner setting’ and ‘individuals’ domain. HCPs noted that they could manage most problems on their own and there were consequently few occasions to consult the cardiovascular expert team (‘tension for change’ construct, see table 1, Q1-Q3). They also mentioned that using the multi-component intervention was not part of their daily routine, making behaviour change as a HCP more difficult and there was no real incentive to change. For example, in one practice BP monitors from the box were rarely used because BP was routinely measured on-site, which was working well for everyone (‘incentive systems’ construct, Q4). In addition, the innovation felt less relevant compared with other tasks, especially since HCPs experience a high workload (‘relative priority’ construct, Q5-Q7). It was mentioned that NPs in particular consulted the expert team, as cardiovascular care is their main focus (‘innovation deliverers’ construct, Q8-10).

**Table 1.** Illustrative quotes accompanying main barriers and facilitators.

| **Domains CFIR** | **Constructs** | **Illustrative quotes** |
| --- | --- | --- |
| Inner setting | Tension for change (barrier) | **Q1 GP1** 'And, I also feel that on a typical day there aren't many moments when you think, oh, I can't figure this out. So the number of questions [towards the expert team], the number of instances will be limited.'<br>**Q2 GP2** 'I had originally expected [that we had to change a lot in people's therapy], because so many people hadn't had a proper check-up for such a long time, or maybe I thought, okay, we should really..., we can still do a lot or at least help think it through.. But actually it wasn't so bad, and that often by just continuing what we were already doing we came to a solution. Yes, I was quite surprised about that.'<br>**Q3 GP5** 'Well, I have literally, I think I probably refer less than 1% per year to an internist for something like that, yes. I am really sure about that. I think maybe less than one or two per year. Very few, I can practically count them on one hand. The last one I referred was that woman we talked about just now and previously. I can't even recall her name.' |
|  | Incentive Systems (barrier) | **Q4 GP1** 'And it is not yet a routine. So yes, altering behaviour [of both patient and care provider], there's just no incentive to change, because we already have that option [measuring blood pressure]. Now suppose that there's no one here [at the practice] who could measure it but there is just a packet ready here for the patient, just pick it up [blood pressure monitor from the box] and you can do it yourself; then it [use of a blood pressure monitor from the box] would become more common, I think. But there's no need for that because it [measuring blood pressure at the practice] is still available.' |
|  | Relative priority (barrier) | **Q5 NP1** 'Yes, and I also think that due to multiple overlapping studies, that it, and because they all started at around the same time, you tend to focus on one thing and tend to forget the other things perhaps?'<br>**Q6 NP2** 'What's the current experience. We are already so incredibly busy. So yes, we also need to pay attention to that.'<br>**Q7 GP1** 'There are really a lot of things on your mind. But at some point they slip down the priority list I think.' |
| Individuals | Implementation deliverer | **Q8 GP5** 'O: Is it mainly the NPs who have been doing this?<br>H: Mainly, of course they spend a larger part of their |
|  |  | *day on this. Yes, I'm mainly involved initially or if the NP can't manage or something, then they call me in again. Yes.'*<br>**Q9 GP4** *"Actually yes, or something comes up when discussing the case, such as whether this might be something we need advice on? But that I actually leave to you [NP]. As far as I know you mainly consult the expert team. And yes, in our case, if it's not embedded in the system, we do it more informally. So maybe a teleconsultation with a nephrologist, but you do that with the expert team.'*<br>**Q10 GP1** *Mainly the NP I think, they asked the most questions.'* |
|  | Implementation leads (barrier) | **Q11 GP1** *'There are 2 categories, so yes, maybe if you really want it to be successful then I guess you would have to stand there every day and say, here, he can take the meter home, etc. But of course that's not possible, just not feasible.'* |
| Implementation process | Teaming/planning (barrier) | **Q12 GP1** *'My biggest mistake in that regard was that it was just me here with you, that the others were not included to help see if it was practical and feasible, because I noticed quite quickly a certain reluctance and that it was quite problematic to hand those things out and it... Just the issue of how to get them back? And, how are you supposed to keep track of people who have handed it in again? You just know that it is already going to be difficult in terms of implementation. Those little practical issues that you run up against.'*<br>**Q13 GP3** *'I need to involve more of my colleagues in the project. No single person is going to shoulder it. That is the first thing you need to do.'* |
| Innovation | Complexity (barrier) | **Q14 GP2** *'We did try to use the blood pressure monitors, but they came back from many patients with the following comment: Too complicated.'*<br>**Q15 GP4** *'No, we tried to use the blood pressure monitors ourselves. We found them quite difficult to set up on your phone and to connect to, and then disconnecting was completely impossible. So we then decided not to use them with patients, because you obviously lose so much time connecting and disconnecting.'*<br>**Q16 GP1** *'No, I think that's a real pity because I had higher expectations myself, and it seems stupid but it's just too complicated. Okay, you can't do anything about that, but the fact is that someone has to come here to* |
|  |  | *pick it up. It's all very practical and then you have to explain how it works and so on; in that case the assistant could just as well have measured blood pressure and that's more or less what actually happened.'* |
|  | Innovation design (facilitator) | **Q17 GP1** *'Yes, that's good. Yes, that's definitely the case. You can really work with the answers they give. It's really quite practical.'*<br>**Q18 NP2** *'Yes, fine. Yes, that we get clear answers that I could do something with.'*<br>**Q19 GP3** *'It's very good, we got very detailed answers.'* |
|  | Relative advantage (facilitator) | **Q20 NP3** *'Well, I think with blood pressure and with medication, when someone is indeed already on a number of things, and you're not sure anymore. You can sometimes suffer from tunnel vision yourself but then you have others who can take a look, that always helps.'*<br>**Q21 GP2** *'We really have had patients where I thought, yes, it is nice to be able to discuss this with someone. But this person doesn't necessarily need to see the internist.'* |

The second main barrier was the insufficient number of individuals in the workplace who could support implementation, which was observed within the ‘individuals’ and ‘implementation process’ domains. HCPs reported that having a dedicated person available to enhance implementation could have helped to maintain priority (‘implementation leads’ construct, Q11). The importance of team involvement was also mentioned (‘teaming’ construct Q12 and Q13).

The third main barrier was that the BP monitors in the toolbox were too complicated to use in practice. This was identified within the ‘complexity’ construct of the ‘innovation’ domain. The BP monitor could only be used with an app, which made it less accessible. Additionally, HCPs stated that BP monitor use had associated difficulties, such as explaining its functionality and managing the collection of BP monitors. As a result it was easier to continue measuring BP in the traditional way – at the general practice (Q14-Q16).

### Facilitators

The most important facilitator was that when HCPs consulted the cardiovascular expert team they found the provided advice helpful, which was assigned to the ‘design’ and ‘relative advantage’ construct of the ‘innovation’ domain. HCPs expressed appreciation for the option to consult the cardiovascular expert team, viewing it as a valuable resource for enhancing patient care and decision making (Q17-Q21).

#### Survey

The DIBQ questionnaire was sent to all participating HCPs and filled out by 15 HCPs (mean age 48 years (sd 8)). Of these HCPs, 60% were GPs and 40% were NPs. ‘Social influences’ and ‘Nature of the behaviour’ received the lowest scores (2.3 and 2.5 respectively, see table 2), indicating that these domains represent barriers for implementation. ‘Social influences’ was defined as ‘those interpersonal processes that can cause individuals to change their thoughts, feelings, or behaviours’, and corresponds with a ‘lack of support for implementation on the work floor’ from the interviews. ‘Nature of the behaviour’ is defined as ‘automaticity’ or ‘habit.’ This suggests that participants did not have interpersonal interactions that sufficiently encouraged them to use the intervention and it therefore did not become a habit, which was in line with the identified barrier from the interviews on ‘a lack of perceived urgency’.

**Table 2.**
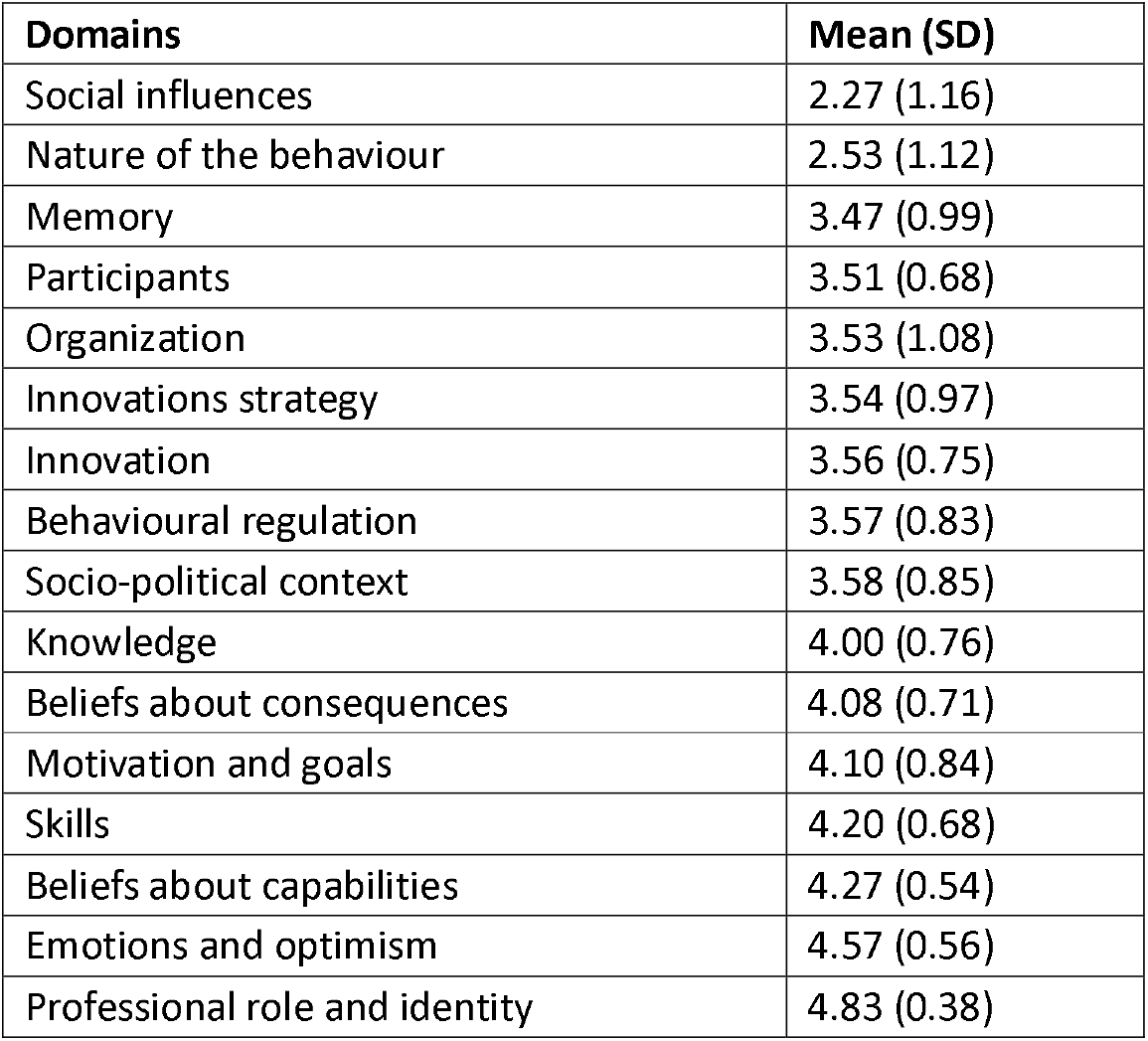
Results of the DIBQ, scale 1-5; the lower the score the higher the barrier on this domain.

| **Domains** | **Mean (SD)** |
| --- | --- |
| Social influences | 2.27 (1.16) |
| Nature of the behaviour | 2.53 (1.12) |
| Memory | 3.47 (0.99) |
| Participants | 3.51 (0.68) |
| Organization | 3.53 (1.08) |
| Innovations strategy | 3.54 (0.97) |
| Innovation | 3.56 (0.75) |
| Behavioural regulation | 3.57 (0.83) |
| Socio-political context | 3.58 (0.85) |
| Knowledge | 4.00 (0.76) |
| Beliefs about consequences | 4.08 (0.71) |
| Motivation and goals | 4.10 (0.84) |
| Skills | 4.20 (0.68) |
| Beliefs about capabilities | 4.27 (0.54) |
| Emotions and optimism | 4.57 (0.56) |
| Professional role and identity | 4.83 (0.38) |

The highest scores were given on the domains ‘Emotions and optimism’ (4.6) and ‘Professional role and identity’ (4.8) (see table 2). This suggests that participants approached the multi-component intervention positively and they considered it their responsibility to use it, which is consistent with the interview findings on the availability of the cardiovascular expert team.

#### Strategies

Based on the described barriers and facilitators in the interviews and survey, we identified 3 strategies to improve implementation. To improve both ‘urgency’ and involve ‘implementation leads’ we selected the ‘identify and prepare champions’ strategy. This could be implemented by selecting a motivated person in every practice who will remind and motivate colleagues to use the innovation. In addition, we selected ‘develop a formal implementation blueprint’, to help keep the innovation a priority and to reduce complexity. The strategy ‘promote adaptability’ was selected for the barrier ‘complexity of BP monitors’ and the observation that NPs made greater use of expert team expertise than GPs. A focus on NPs instead of GPs might also increase adoption of the intervention. As HCPs often mentioned that patients were willing to buy their own BP monitor, we anticipate that implementation will improve if patients can choose their own BP monitor.

#### Consultations with the cardiovascular expert team

During the study, a total of 33 consultations involving different patients were requested with the cardiovascular expert team (table 3). Advice to continue treatment in primary care was given in 31 of these cases. In the absence of the cardiovascular expert team consultation, 45.2% (14/31) would have resulted in a consultation with a medical specialist and 35.5% (11/31) in referral to secondary care.

**Table 3.** Number of referrals to the cardiovascular expert team during the study period.

|  | Advice from cardiovascular expert team |  | Total<br>(n=33) |
| --- | --- | --- | --- |
|  | Treatment in primary<br>care (n=31) | Referral to secondary<br>care (n=2) |  |
| **Subject** |  |  |  |
| Hypercholesterolemia | 6 (19.4%) | 0 (0%) | 6 (18.2%) |
| Hypertension | 16 (51.6%) | 1 (50.0%) | 17 (51.5%) |
| Other | 9 (29.0%) | 1 (50.0%) | 10 (30.3%) |
| **What would the healthcare**<br>**professional have done otherwise?** |  |  |  |
| Call medical specialist for advice | 14 (45.2%) | 1 (50.0%) | 15 (45.5%) |
| Continue treatment in primary care | 4 (12.5%) | 0 (0%) | 4 (12.1%) |
| Referral to secondary care | 11 (35.5%) | 1 (50.0%) | 12 (36.4%) |
| Missing | 2 (6.5%) | 0 (0%) | 2 (6.1%) |
| **Answered by** |  |  |  |
| General practitioner | 9 (29.0%) | 0 (0%) | 9 (27.3%) |
| Nurse practitioner | 13 (41.9%) | 2 (100%) | 15 (45.5%) |
| Nurse practitioner + General<br>practitioner | 8 (25.8%) | 0 (0%) | 8 (24.2%) |
| Nurse practitioner + Medical<br>Specialist | 1 (3.2%) | 0 (0%) | 1 (3.0%) |

#### Patient-related characteristics

From the 13 practices 591 patients were invited to participate, of whom 176 (29.8%) gave informed consent and 165 completed the whole study. The average age of participants at baseline was 70 years and 47% was female. The average number of measurements per patient during follow-up was 1.6 (min 0, max 7). Mean systolic BP was 158 (15 sd) mmHg before the intervention and 148 (21 sd) mmHg during follow-up. The decrease in systolic BP was higher in the high SEP group (from 159 to 148 mmHg) than in the low SEP group (156 to 151 mmHg). Mean diastolic BP decreased from 87 (11 sd) mmHg before implementation to 85 (13 sd) mmHg during follow-up. The number of prescribed BP-lowering medications changed from 2.8 (1.3 sd) at the start of the study to 3.0 (1.3 sd) during follow-up. See table 4 for full participant characteristics and changes in eGFR, BMI and LDL. Due to the use of routine care data approximately 10%-60% of the data were missing, as measurements were only taken when clinically appropriate. However, an additional analysis focusing on the complete cases showed descriptive statistics comparable to those reported for the full sample (see Supplementary Table 1).

**Table 4.**
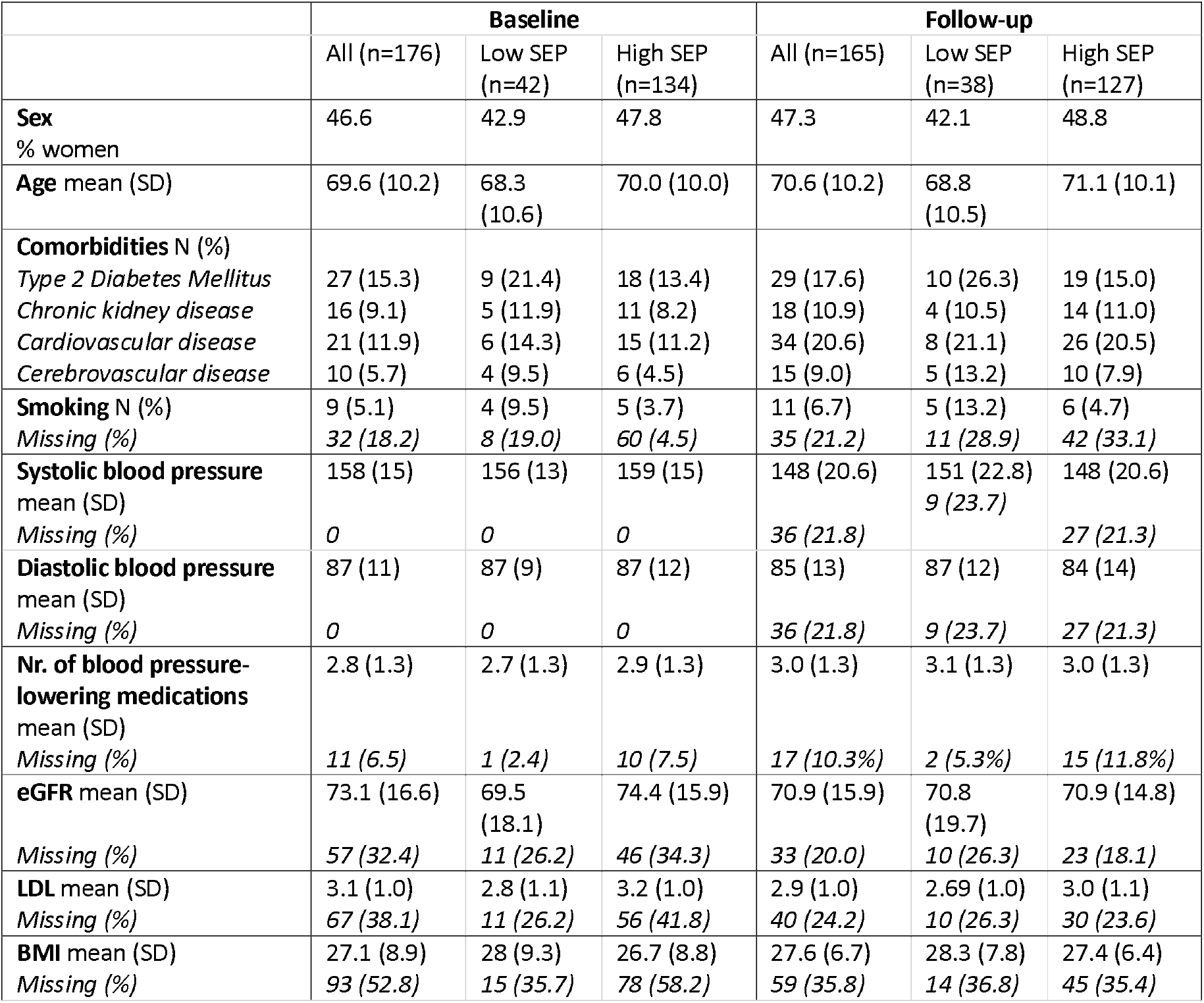
Characteristics of participants with hypertension. Due to the use of routine data not all values were available for all participants, as these were only measured when clinically relevant.

## Discussion

In this implementation study, we aimed to assess the adoption and feasibility of a multi-component intervention designed to ensure that patients receive appropriate care, reduce avoidable referrals and support HCPs in hypertension management. In addition, we evaluated potential changes in health outcomes. Our findings showed that although HCPs were generally open to innovation adoption, urgency was sometimes lacking. While the cardiovascular expert team was well-received and considered practical, intervention tools were considered challenging to use. We identified three evidence-based and expert-recommended strategies to improve future implementation: ‘identify and prepare champions’, ‘develop a formal implementation blueprint’ and ‘promote adaptability’. Results of BP measurements were also promising, especially among individuals in the low social deprivation group.

In its current form, intervention adoption by HCPs was only moderate. A barrier to adoption was a lack of perceived urgency to consult the cardiovascular expert team by GPs, as they already had existing workflows that they considered effective. This low sense of urgency was somewhat unexpected given that uncontrolled hypertension is a major contributor to cardiovascular morbidity and mortality(18). This discrepancy may be due to competing clinical priorities, limited awareness of the intervention’s potential benefits, or confidence in current management strategies. Furthermore, during the course of the study hospitals in the region started to use a similar teleconsultation service that appeared at the top of the list when opening the referral system, while the cardiovascular expert team could only be found via the search option. This might have contributed to the relatively low engagement with the cardiovascular expert team, and also aligns with previous literature on implementation determinants, which found that the relationship of an innovation to existing solutions plays an important role in its uptake. People will only accept an innovation if it satisfies their needs to a greater extent than an existing solution, and if the benefit is worth the financial and non-financial effort, such as changing ingrained habits (19). In this study the overlap with existing services and the lack of perceived added value likely reduced motivation for adoption. Other barriers were difficulties incorporating the intervention into the daily workflow and the presence of competing priorities. This finding is in line with prior studies concerning digital health implementation, which found clinician-related barriers concerning lack of workflow integration and the additional time required to use new innovations alongside routine tasks (20, 21). Despite these barriers the cardiovascular expert team was viewed as practical, suggesting that acceptability was not an issue, although workflow alignment could be improved.

While the cardiovascular expert team was considered feasible for HCPs, the digital tools in the toolbox were often perceived as too complex. HCPs frequently mentioned that despite initial expectations that an app-supported BP monitor would be helpful, most patients found the app difficult to use. Previous studies have also highlighted barriers faced by HCPs when using digital apps, such as insufficient technical support (22). Limited health literacy was also mentioned as a barrier in relation to digital apps (23), although in our study even individuals with good health literacy experienced difficulties using the BP monitor and app. While a previous study reported that the BP monitors were implemented successfully, this was accompanied by substantial technological support for each patient (24), suggesting that the technology itself may be too complex for effective use in a primary care setting without technical support.

Our findings suggest a need for additional implementation strategies. Identifying and preparing champions, individuals who actively promote and support the intervention, can help to ensure priority and visibility of new interventions. Regardless of who takes on the role of health champion, commitment to the intervention is more important than the person’s profession, as shown in previous research on weight loss interventions in primary care(25). In line with this, successful implementation of an innovation needs ‘promotors’, key persons who make adoption happen (19, 26, 27) and are willing to support an innovation through special commitment beyond daily responsibilities. In the context of our study both NPs and GPs could play this role, depending on their level of involvement and motivation in cardiovascular care. Additionally, practice managers could also act as ‘promotors’, given their knowledge of operational workflows and team coordination.

Although we designed our intervention (the cardiovascular expert team and toolbox) based on perceived barriers in hypertension management by GPs and medical specialists (9), we did not use a formal implementation plan. A ‘formal implementation blueprint’, including context-sensitive multiple strategies from multiple domains, may help to give structure to the new workflow and can serve as a reminder for HCPs. This plan can also be updated and guide implementation over time (28). These strategies could therefore help stimulate engagement with the cardiovascular expert team. For instance, the strategy ‘promoting adaptability’ could be applied to modify the type of BP monitor used in the innovation. Since home BP monitoring is known to play a crucial role in managing hypertension (29-31), using a home BP monitor remains beneficial for patients. However, it may be more effective to use a simpler device.

Another important finding from interviews was the revealed differences in perceived relevance between professional roles. GPs stated that cardiovascular risk management was not their main focus during the day, as they have many other tasks. In contrast, cardiovascular care is the main focus for NP’s, so for them consultation with the cardiovascular expert team might potentially be more relevant. In the Netherlands, the number of NPs working in general practices has increased rapidly over the past two decades; by 2023, 94% of general practices had a NP working in the practice, 38% of NPs had daily discussions with the GP, and 26% consulted the GP multiple times a week (32). Focusing implementation of the cardiovascular expert team more towards NPs instead of GPs would likely enhance the intervention’s impact, as well as save time and reduce GPs workloads. However, adapting the intervention to place greater focus on NPs should be explored to ensure it supports collaboration between NPs and GPs rather than reinforcing role separation, as GPs should remain actively involved in their patient’s care.

This study has several limitations. First, since routine clinical data were collected over the study period, data completeness was variable. Nonetheless, complete case analysis results were mainly consistent with the overall analysis, indicating that missing data are unlikely to have substantially biased the findings. Second, while we observed improvement in BP there is potential for regression to the mean, since we selected patients with elevated BP at baseline and did not include a formal control group. However, regarding both missing data and potential for regression to the mean, the study was primarily focused on implementation and not evaluation of changes in BP. Nevertheless, it is noteworthy that we did not observe an increase in BP during the study period, suggesting the absence of a negative impact on BP outcomes. Lastly, due to privacy constraints it was not possible to link specific consultations to patient-level BP data.

Strengths of the study include the theory-informed approach in which barriers, facilitators and accompanying strategies were identified. The CFIR and TDF (DIBQ) are well-known and often used in healthcare settings, allowing for a systematic analysis of results. In addition, the diversity of participating practices, from both high and low SEP neighbourhoods, enhances the generalizability of our findings. Finally, this study contributes to research on interdisciplinary collaboration in hypertension care. By evaluating an innovation that combines both primary and secondary care expertise in the form of a cardiovascular expert team, we showed how such collaborative approaches can be optimized to fit existing healthcare settings.

Overall, the study emphasized the importance of structured implementation strategies to optimize hypertension care interventions. Primary and secondary care working together in the form of a cardiovascular expert team, teleconsultation and self-management tools are valuable ways to improve hypertension management. To ensure adoption and feasibility, future implementation efforts should focus on engaging NP’s as key users of the cardiovascular expert team and applying strategies such as formal implementation plan and the identification of champions.

## Supporting information

Supplementary Box 1

Supplementary Box 2

Supplementary Box 3

## Data Availability

The data that support the findings of this study are available from ELAN but restrictions apply to the availability of these data, which were used under license for the current study, and so are not publicly available. Data are however available from the authors upon reasonable request and with permission of ELAN.

## Declarations

### Ethics approval and consent to participate

The department’s science committee positively assessed the current study’s quality and feasibility (#WSC-2020-30/SP) and the Medical Ethics Review Board of Leiden University Medical Center confirmed that the Medical Research Involving Human Subjects Act (WMO in Dutch) does not apply to the current study according to Dutch standards (Protocol number: P20-092).

### Authors’ contributions

SG, RV, SB, HV, MN, GR and MB contributed to the design of the study. SG conducted the interviews. SG, AE and RV contributed to the qualitative analysis, SG and JB did the quantitative analyses. SG drafted the manuscript. RV, HV, SB, JB, AE, MN, GR and MB helped drafting and revising the manuscript. All authors read and approved the final manuscript.

## Acknowledgements

We thank J.M. Huijg and M.R. Crone for kindly providing the DIBQ utilized in this research.

