## Supplementary figures and images for "Optimized hypertension care for people with high blood pressure by improved integrated care and self-management tools: a mixed-methods study"

### Supplementary Box 1

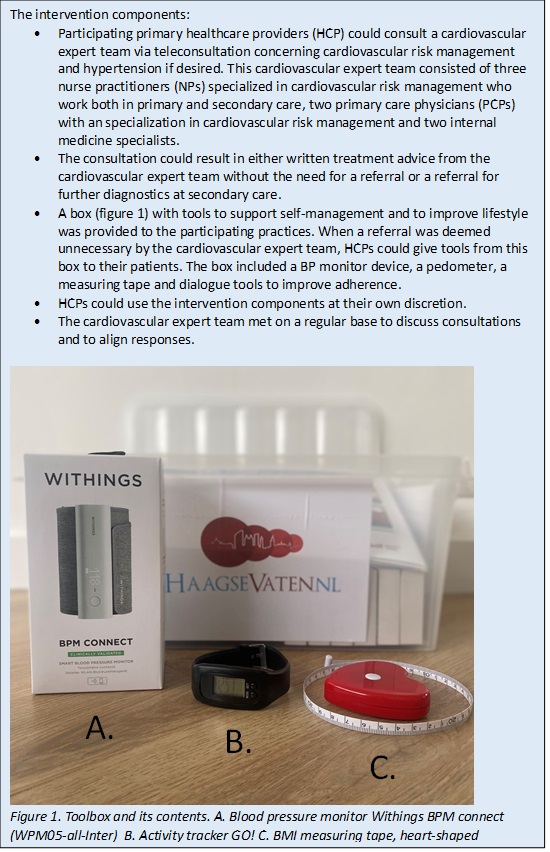
