## Supplementary Box 2 for "Optimized hypertension care for people with high blood pressure by improved integrated care and self-management tools: a mixed-methods study"

Supplementary Box 2: Interview guide

1. How have you organized the intervention?
2. Which components of the intervention have you utilized?
   a. What did you think of them?
   b. To whom/what type of patient did you provide this?
3. What is the reason you haven't used [particular components of the intervention]?
   a. Do you see added value in using [particular components of the intervention]?
      i. If yes, what would make you use [particular components of the intervention] more?
      ii. If no, how could we improve/modify it?
