## Supplementary Box 3 for "Optimized hypertension care for people with high blood pressure by improved integrated care and self-management tools: a mixed-methods study"

**Supplementary Box 3: DIBQ Questionnaire**

Answer options for each quote:

Totally disagree	1	2	3	4	5	Totally agree

1. It is clear to me which activities I need to do and in which order I need to do them to make a referal to the expert team following the guidelines. [innovation]
2. The procedure of making a referral to the expert team is well-constructed (content wise). [innovation]
3. Making a referral to the expert team is compatible with how I am accustomed to work. [innovation]
4. The expert team offers all information and materials that are necessary to make a referral following the guidelines. [innovation]
5. It is possible to tailor a referral to the expert team to participants’ individual characteristics and needs. [innovation]
6. The effects of making a referral to the expert team are clearly visible to me. [Innovation]
7. Making a referral to the expert team gives me a lot of benefits. [innovation]
8. I have sufficient knowledge to make a referral to the expert team following the guidelines. [knowledge]
9. I have sufficient skills to make a referral to the expert team following the guidelines.[Skills]
10. I think that as a professional it is my job to treat people with hypertension. [professional role and identity]
11. I think that as a professional is it my job to ask advice when I am not sure about the treatment of a patient with hypertension. [professional role and identity]
12. I am motivated to make use of the expert team. (Motivation and Goals]
13. If I make a referral to the expert team this will lead to improvement of my knowledge about cardiovascular risk management. [Beliefs about consequences]
14. If I make a referral to the expert team this will lead to the right treatment for my patients. [Beliefs about consequences]
15. The expert team improves the collaboration between primary care and secondary care. [Beliefs about consequences]
16. If I make a referral to the expert team this will lead to less referrals to secondary care.
17. I am confident that I can treat people with hypertension. Beliefs about capabilities]
18. I ask advice when I am not sure about the treatment of someone with hypertension. [Beliefs about capabilities]
19. I am confident I can make a referral to the expert team even when I encounter barriers. [Beliefs about capabilities]
20. I feel good when I make a referral to the expert team if I am not sure about the treatment of someone with hypertension. [emotions and optimism]
21. *I do not like to make a referral to the expert team if I am not sure about the treatment of someone with hypertension. [emotions and optimism]*
22. *Other work tasks I need to do interfere with making a referral to the expert team. [Motivation and goals]*
23. I have clear plans of how I make a referral to the expert team following the guidelines. [Behavioural regulation]
24. I have clear plans of how I make a referral to the expert team following the guidelines when I encounter barriers. [Behavioural regulation]
25. I can easily remember that I can make a referral to the expert team. [Memory]
26. Making a referral to the expert team is something I have made my own/that has become habitual for me. [Nature of the behaviors]
27. There are sufficient financial resources to make a referral to the expert team. [Socio-political context]
28. I have sufficient time to make a referral to the expert team. [Socio-political context]
29. I experience the collaboration with the expert team as positive. [Socio-political context]
30. The expert team is well coordinated. [Socio-political context]
31. In my organization, formal arrangements are made with regard to making a referral to the expert team. [Organization]
32. In my organization, there are sufficient facilities to make a referral to the expert team following the guidelines. [Organization]
33. *In my organization, other changes interfere with making a referral to the expert team. [Organization]*
34. It is expected from me that I make referrals to the expert team following the guidelines. [social influences]
35. I can count on sufficient support from people when I make a referral to the expert team. (e.g. from colleagues, management, others involved) [Social influences]
36. In my organization, there is sufficient influx of patients to make referrals to the expert team. [participants]
37. In general patients are positive about the expert team. [participants]
38. Participants are motivated to participate in the advice of the expert team. [participants]
39. I would like to have (more) information on how to make a referral to the expert team following the guidelines. [innovation strategy]
40. I would like to have (more) training on how to make a referral to the expert team following the guidelines. [innovation strategy]
41. I would like to have (more) assistance on how to make a referral to the expert team following the guidelines. [innovation strategy]
42. I get informed regularly about the progress of the expert team. [innovation strategy]
43. I get sufficient financial reimbursement when I make a referral to the expert team. [innovation strategy]
