## Supplementary Table 1 for "Optimized hypertension care for people with high blood pressure by improved integrated care and self-management tools: a mixed-methods study"

**Table S1: Complete case analysis for blood pressure.**

|  | **Baseline complete cases** | | | **Follow-up** | | |
| --- | --- | --- | --- | --- | --- | --- |
|  | All (n=129) | Low SEP (n=29) | High SEP (n=100) | All (n=129) | Low SEP (n=29) | High SEP (n=100) |
| **Sex**  % women | 48.1 | 37.9 | 51.0 | 47.7 | 37.9 | 50.5 |
| **Age** mean (SD) | 70.0 (10.0) | 68.9 (8.6) | 70.4 (10.3) | 70.6 (10.2) | 68.8 (10.5) | 71.1 (10.1) |
| **Mean Systolic blood pressure** (SD) | 158 (15.2) | 156 (12.9) | 158 (15.9) | 148 (20.6) | 151 (22.8) | 148 (20.6) |
| **Mean Diastolic blood pressure** (SD) | 87 (11) | 89 (9) | 86 (11) | 85 (13) | 87 (12) | 84 (14) |
| **Nr. of blood pressure-lowering medications**  mean (SD)  *Missing (%)* | 2.8 (1.3)  *11 (6.5)* | 2.7 (1.3)  *1 (2.4)* | 2.9 (1.3)  *10 (7.5)* | 3.0 (1.3)  *17 (10.3%)* | 3.1 (1.3)  *2 (5.3%)* | 3.0 (1.3)  *15 (11.8%)* |
